# OGTT Early C-Peptide Index for Clamp-Measured Acute β-Cell Secretory Capacity

**DOI:** 10.64898/2026.10.05.26364738

**Authors:** Ren Zhang

**Affiliations:** Center for Molecular Medicine and Genetics. Wayne State University School of Medicine, Detroit, MI 48202, USA

**Keywords:** Oral glucose tolerance test, C-peptide, hyperglycemic clamp, insulin secretion

## Abstract

Acute glucose-stimulated β-cell secretion is a key dimension of β-cell function, but the hyperglycemic clamp used to assess it is technically demanding and resource-intensive, limiting its use at scale. We aimed to develop and validate the oral glucose tolerance test (OGTT) Early C-peptide (OEC) Index for estimating clamp-derived acute C-peptide response to glucose (ACPRg). We defined OEC as the 0–60-min incremental C-peptide area under the curve (AUC) normalized to the geometric mean of incremental and total glucose AUCs, with a fasting-glucose correction of 100/G₀. OEC was evaluated in paired OGTT–clamp data from RISE Adult Medication (baseline n=257; internal validation) and BetaFat (baseline n=70; external validation). Predictive R² and Pearson r for log-ACPRg were assessed using participant-level 5-fold cross-validation in RISE and external validation without refitting in BetaFat, with the C-peptidogenic, insulinogenic, and CIR30 indices as comparators. In RISE Adult, cross-validated R² for OEC was 0.47, 0.61, and 0.54 at baseline, 12, and 15 months, respectively, with corresponding Pearson r of 0.69, 0.78, and 0.74; nested cross-validation showed only modest attenuation. Applied without refitting in BetaFat, OEC yielded R² of 0.55, 0.41, and 0.39 at baseline, 12, and 24 months, with corresponding r of 0.76, 0.70, and 0.64. OEC had higher predictive R² point estimates than all three comparators at every visit in both cohorts. OEC provides a simple, transparent OGTT-based estimate of acute β-cell secretory capacity that retained predictive performance in BetaFat and may broaden access to physiological assessment of early β-cell function.

## Introduction

Diabetes and prediabetes affect a substantial and growing proportion of the population worldwide [1]. Type 2 diabetes develops when pancreatic β-cell insulin secretion is insufficient to compensate for insulin resistance [2]. In a longitudinal human study, the acute insulin response to glucose declined as participants progressed from normal glucose tolerance to impaired glucose tolerance and diabetes; a multicenter study also found substantial first-phase secretory defects in people with impaired glucose tolerance and early diabetes [3, 4]. Loss of the early insulin response can contribute to postprandial hyperglycemia by delaying suppression of endogenous glucose production [5, 6]. In individuals with impaired glucose tolerance, absence of a first insulin peak during hyperglycemic clamp was a strong predictor of progression to type 2 diabetes [7]. Because C-peptide is secreted equimolarly with insulin but is not subject to hepatic first-pass extraction, it provides a useful measure of pancreatic β-cell secretion. Acute glucose-stimulated β-cell secretion is therefore a key dimension of β-cell function to characterize in dysglycemia.

The hyperglycemic clamp assesses β-cell responsiveness by raising plasma glucose to a predefined hyperglycemic level with intravenous glucose and maintaining it there while insulin and C-peptide are measured serially [8, 9]. Its controlled glucose stimulus and temporal resolution allow early and sustained secretory responses to be assessed, making it a physiological reference method for glucose-stimulated β-cell function. However, the procedure requires intravenous access, frequent blood sampling and glucose measurements, and skilled adjustment of the glucose infusion to maintain the target. These demands make the clamp time- and resource-intensive and difficult to use at scale [9].

The oral glucose tolerance test (OGTT) is an established test for diagnosing diabetes and identifying impaired glucose tolerance. It is substantially simpler and less resource-intensive to perform than a hyperglycemic clamp. When insulin or C-peptide is measured alongside glucose, OGTT responses can also be used to characterize β-cell function. OGTT-derived β-cell indices include insulinogenic indices based on insulin or C-peptide and corrected insulin response [10–12]. Related OGTT measures have been examined against IVGTT or clamp responses and for prediction of dysglycemia [13–16]. However, accurately estimating clamp-measured acute C-peptide response to glucose (ACPRg) from the OGTT remains challenging. One physiological consideration in developing such a measure is that fasting and post-load glycemia reflect partly distinct aspects of glucose homeostasis. In a large cohort without diabetes, fasting plasma glucose and mean post-load glucose increment were weakly associated (standardized β = 0.12), and only 23% of participants were in the same quintile for both measures [17]. This distinction motivated us to evaluate a simple OGTT-derived index that accounts for fasting glycemia and relates early C-peptide secretion to both the glucose increment above fasting and total glucose exposure. We therefore aimed to develop and internally validate the OGTT Early C-peptide (OEC) Index for estimating clamp-derived ACPRg in the RISE Adult Medication Study, compare its performance with established OGTT-derived indices, and externally validate it in the independent BetaFat cohort.

## Materials and methods

### Study populations and data sources

The present study was a secondary analysis of participant-level data from two adult studies of the Restoring Insulin Secretion (RISE) Consortium obtained through the NIDDK Central Repository: the RISE Adult Medication Study and the Beta Cell Restoration Through Fat Mitigation (BetaFat) Study. The Adult Medication Study included adults with impaired glucose tolerance or recently diagnosed type 2 diabetes who underwent oral glucose tolerance testing (OGTT) and hyperglycemic clamp assessments at baseline, 12 months, and 15 months [18, 19]. BetaFat included a separate adult RISE population evaluated at baseline, 12 months, and 24 months [18, 20]. The Adult Medication Study was used for development and internal validation of OEC, whereas BetaFat was used for external validation. Detailed study designs, eligibility criteria, and metabolic testing protocols have been reported previously [18–20].

The baseline analytic populations comprised 257 participants in the Adult Medication Study and 70 participants in BetaFat (Table 1). Corresponding analytic sample sizes were 220 and 207 at 12 and 15 months in the Adult Medication Study and 56 and 53 at 12 and 24 months in BetaFat, respectively. OGTT glucose, insulin, and C-peptide measurements were used to calculate OEC and the comparator indices. Because no sample was obtained at exactly 0 min, fasting glucose, C-peptide, and insulin were defined as the mean of the −10- and −5-min measurements. The reference outcome was the clamp-derived acute C-peptide response to glucose (ACPRg), calculated as the mean incremental C-peptide response above baseline from samples obtained at 2, 4, 6, 8, and 10 min after intravenous glucose administration [21]. Analyses were restricted to participants with positive ACPRg and complete measurements required for calculation of OEC and all comparator indices at the corresponding visit. Missing values were not imputed, and all head-to-head comparisons were performed in the same complete-case sample within each visit. The present study was approved by the Wayne State University Institutional Review Board.

**Table 1.** Baseline characteristics of the analytic populations in the RISE Adult Medication Study and BetaFat Study.

| Characteristic | RISE Adult Medication Study | BetaFat Study |
| --- | --- | --- |
| Participants, n | 257 | 70 |
| Female, n (%) | 110 (42.8) | 56 (80.0) |
| BMI, kg/m<sup>2</sup> | 35.0 ± 5.7 | 35.6 ± 2.9 |
| HbA1c, % | 5.75 ± 0.39 | 5.83 ± 0.41 |
| Fasting glucose, mg/dL | 110.6 ± 10.9 | 108.8 ± 11.1 |
| Fasting C-peptide, ng/mL | 3.69 ± 1.29 | 3.59 ± 1.03 |
| ACPRg, ng/mL | 1.83 [1.00–3.10] | 1.72 [1.02–3.07] |

**Note.** Data are mean ± SD, n (%), or median [IQR]. Fasting glucose and fasting C-peptide were calculated as the mean of the -10- and -5-min OGTT measurements. ACPRg denotes the acute C-peptide response to glucose measured during the hyperglycemic clamp.

### Development and definition of the OEC index

The OEC (OGTT Early C-peptide) index was developed as a physiology-guided measure of early β-cell secretory response. Candidate formulations combined integrated early C-peptide secretion with complementary measures of the glucose stimulus and fasting glycemia. The 0–60-min interval was retained because extending the response window beyond 60 min did not materially improve prediction.

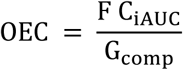

where F = 100/G0 is a dimensionless fasting-glucose correction normalized to 100 mg/dL, C_iAUC_ is the incremental C-peptide area from 0 to 60 min, and G_comp_ is the composite glucose exposure defined as

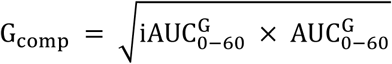

Incremental AUCs were calculated relative to the fasting concentration by trapezoidal integration using the fasting, 30-min, and 60-min measurements, with negative incremental excursions set to zero. Total glucose AUC was calculated over the same interval by the trapezoidal rule. OEC therefore integrates early C-peptide secretion with incremental and total glucose exposure and fasting glycemia (Figure 1). No empirically fitted coefficients enter the index.

**Figure 1.**
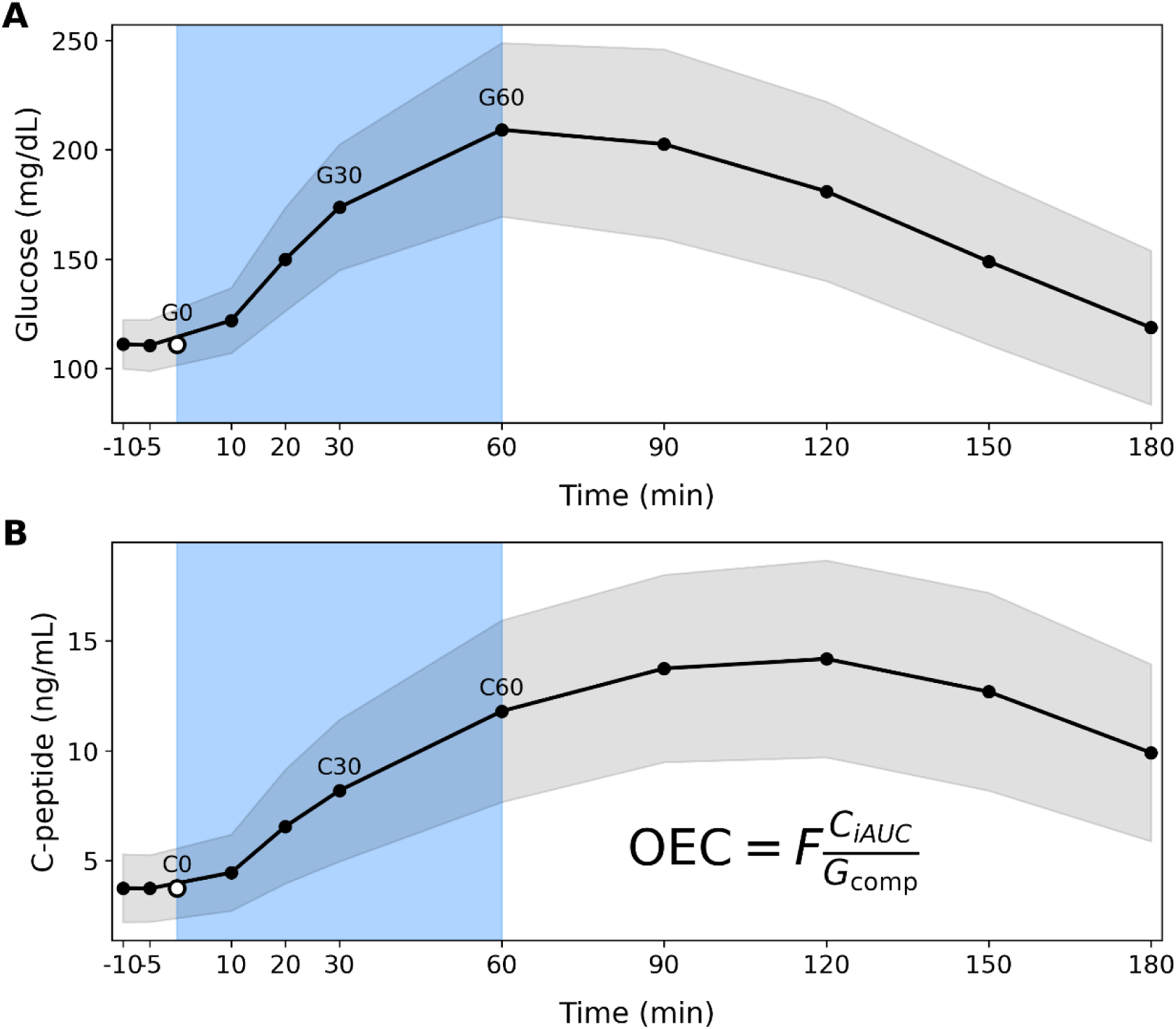
OGTT profiles and definition of the OEC index. Mean glucose. (A) and C-peptide **(B)** values (black circles) with ±1 SD (gray shading) during the baseline oral glucose tolerance test in the RISE Adult Medication Study. The shaded 0–60 min interval identifies the measurements used for OEC calculation. G0 and C0 (open circles) represent fasting values calculated as the mean of the −10- and −5-min measurements; G30, G60, C30, and C60 are measured values at 30 and 60 min. OEC integrates the early incremental C-peptide response with incremental and total glucose exposure and fasting glycemia.

For direct calculation from the fasting, 30-min, and 60-min measurements, the expression can be rewritten as

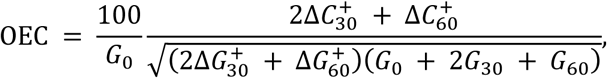

where

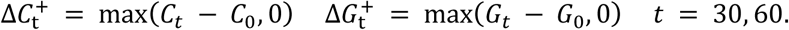

Here, *C_t_* and *G_t_* denote measured C-peptide and glucose concentrations at time t, respectively, and *C*_0_ and *G*_0_ denote their fasting concentrations. The superscript + indicates that negative increments are set to zero. Glucose is expressed in mg/dL and C-peptide in ng/mL. The coefficients arise from trapezoidal integration over the two equal 30-min intervals.

### Comparator indices

OEC was compared with three established OGTT-derived indices of early β-cell response: the C-peptidogenic index, insulinogenic index, and corrected insulin response at 30 min (CIR30) [10–12].

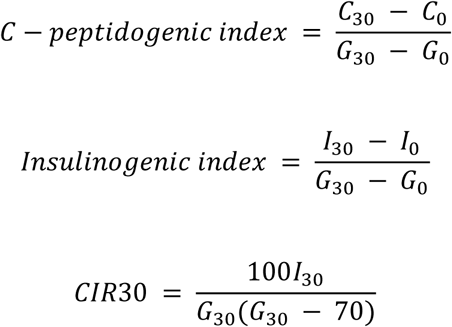

For all indices, G0, C0, and I0 were defined as the mean of the −10- and −5-min measurements, as described above. Glucose was expressed in mg/dL and insulin in μU/mL. Only finite positive index values were included in log-scale prediction analyses.

### Validation and statistical analysis

Internal validation was performed in the RISE Adult Medication Study using participant-level 5-fold cross-validation. Participants, rather than individual visits, were assigned to folds so that all observations from a given participant remained in the same fold. At each visit, ordinary least-squares regression of log-transformed ACPRg on the log-transformed index was fitted using the training folds and applied to the held-out fold. OEC and comparator indices were evaluated using the same complete-case sample within each visit. A nested 5-fold cross-validation analysis, in which index selection was repeated within each outer training set, was additionally performed to assess optimism related to formula development.

For external validation, regression coefficients for OEC and each comparator were estimated using baseline data from the RISE Adult Medication Study and then fixed. The resulting models were applied without recalibration or refitting to BetaFat participants at baseline, 12 months, and 24 months. Predictive performance was quantified by predictive R² and the Pearson correlation coefficient between observed and predicted log-transformed ACPRg. Predictive R² was calculated as

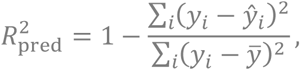

Where *y_i_* and ŷ*_i_* are the observed and predicted log-transformed ACPRg values, respectively, and ȳ is the mean observed log-transformed ACPRg. Ninety-five percent confidence intervals were estimated by participant-level bootstrap resampling with 10,000 replicates. Baseline continuous variables were summarized as mean ± SD, except for ACPRg, which was summarized by the median and interquartile range; categorical variables were summarized as number and percentage. Analyses used complete cases, with no imputation of missing values. Analyses were conducted in Python using NumPy, pandas, and scikit-learn. Figures were generated using Matplotlib.

## Results

### Development and initial evaluation of OEC

The design of OEC was based on a simple physiological premise: an OGTT surrogate of clamp-derived acute β-cell secretory capacity should quantify secretion in relation to the glucose stimulus that elicits it. During a hyperglycemic clamp, the rapid rise in plasma glucose provides a relatively standardized stimulus, and the resulting early increase in C-peptide reflects acute β-cell secretion. During an OGTT, however, both the magnitude and temporal profile of the glucose stimulus vary among individuals, making the C-peptide response difficult to interpret independently of concurrent glucose exposure. Glucose concentrations rose rapidly during the first 60 min of the OGTT before declining thereafter (Fig. 1A), while C-peptide increased progressively over the same early interval (Fig. 1B). OEC was therefore constructed to relate integrated early C-peptide secretion to both incremental and total glucose exposure, with additional adjustment for fasting glycemia, yielding a simple coefficient-free formulation.

The 0–60-min OEC formulation was initially evaluated in the RISE Adult Medication Study using clamp-derived ACPRg as the reference measure. In the complete-case samples, the apparent R² for log-transformed ACPRg was 0.474 at baseline, 0.608 at 12 months, and 0.555 at 15 months, with corresponding Pearson correlations of 0.689, 0.780, and 0.745. Thus, OEC accounted for approximately 47%–61% of the variation in log-transformed ACPRg across the three assessments.

The choice of the early response window was examined by extending both the C-peptide and glucose-exposure windows while retaining the same index architecture. At baseline, cross-validated R² decreased from 0.469 with the 60-min window to 0.445, 0.422, 0.398, and 0.380 with 90-, 120-, 150-, and 180-min windows, respectively. Similar reductions were observed at 12 and 15 months. Across the three visits, mean R² declined from 0.533 at 60 min to 0.457 at 180 min, accompanied by a reduction in mean Pearson r from 0.729 to 0.675. Thus, inclusion of later OGTT measurements provided no improvement in prediction of ACPRg, supporting the 0–60-min interval for the final OEC formulation.

### Internal validation and robustness of OEC

The initial analysis quantified the relationship between OEC and ACPRg within the RISE Adult Medication Study, but performance estimated in the same data used to fit the relationship may overestimate performance in new participants. We therefore used standard participant-level 5-fold cross-validation to assess performance in held-out participants. Participants were partitioned into five mutually exclusive folds; the regression model was fitted in four folds and applied to the remaining fold, with the process repeated until each participant had an out-of-fold prediction. All observations from a given participant were retained in the same fold to prevent information leakage between training and test sets.

OEC retained substantial predictive performance across all three assessments. Cross-validated R² was 0.469 at baseline, 0.607 at 12 months, and 0.544 at 15 months (Fig. 2A; Table 2).

**Figure 2.**
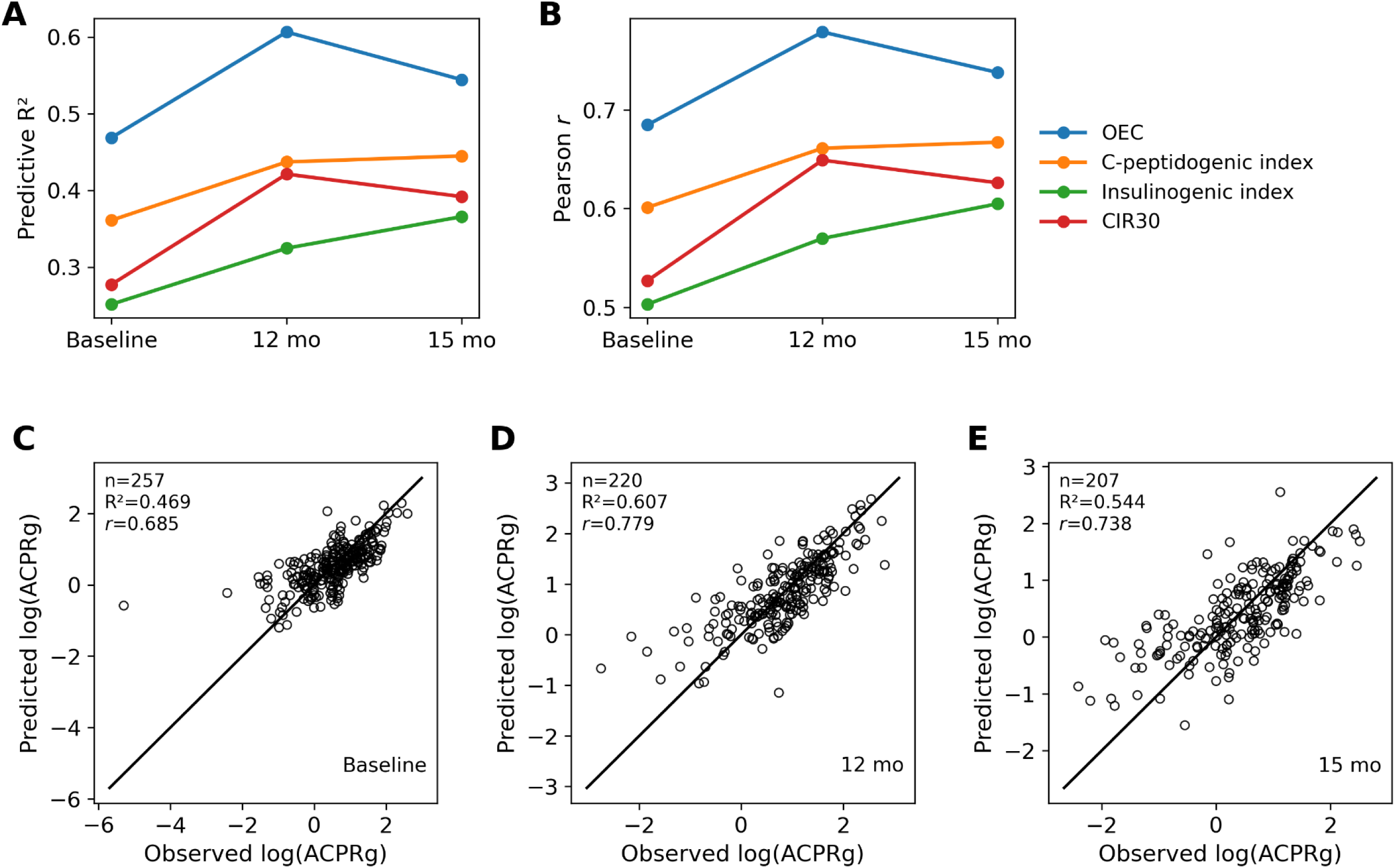
Internal validation of OEC in the RISE Adult Medication Study. Predictive R² **(A)** and Pearson r **(B)** for OEC, the C-peptidogenic index, insulinogenic index, and corrected insulin response at 30 min (CIR30) at baseline, 12 months, and 15 months. Performance was assessed by participant-level 5-fold cross-validation. Observed and out-of-fold predicted log(ACPRg) values for OEC are shown at baseline **(C)**, 12 months **(D)**, and 15 months **(E)**. Individual participants are shown as open circles; the diagonal line represents identity. Sample size, R², and Pearson r are reported in each panel.

**Table 2.** Internal validation of OEC and conventional OGTT indices in the RISE Adult Medication Study.

| Visit | n | Index | 5-fold predictive R<sup>2</sup> (95% CI) | Pearson r (95% CI) |
| --- | --- | --- | --- | --- |
| Baseline | 257 | **OEC** | **0.469 (0.383 - 0.558)** | **0.685 (0.625 - 0.750)** |
|  |  | C-peptidogenic index | 0.361 (0.211 - 0.515) | 0.601 (0.476 - 0.723) |
|  |  | Insulinogenic index | 0.251 (0.108 - 0.400) | 0.503 (0.359 - 0.638) |
|  |  | CIR30 | 0.277 (0.148 - 0.411) | 0.527 (0.401 - 0.647) |
| 12 months | 220 | **OEC** | **0.607 (0.511 - 0.681)** | **0.779 (0.721 - 0.828)** |
|  |  | C-peptidogenic index | 0.437 (0.327 - 0.526) | 0.661 (0.584 - 0.730) |
|  |  | Insulinogenic index | 0.325 (0.232 - 0.403) | 0.570 (0.497 - 0.646) |
|  |  | CIR30 | 0.421 (0.327 - 0.501) | 0.649 (0.580 - 0.712) |
| 15 months | 207 | **OEC** | **0.544 (0.439 - 0.628)** | **0.738 (0.673 - 0.797)** |
|  |  | C-peptidogenic index | 0.445 (0.332 - 0.535) | 0.667 (0.591 - 0.737) |
|  |  | Insulinogenic index | 0.366 (0.249 - 0.463) | 0.605 (0.515 - 0.686) |
|  |  | CIR30 | 0.392 (0.289 - 0.479) | 0.626 (0.550 - 0.697) |

Note: Predictive R<sup>2</sup> and Pearson r were calculated from out-of-fold predictions generated by participant-level 5-fold cross-validation, with the same participants used for all indices within each visit. 95% confidence intervals were estimated by participant-level bootstrap resampling (10,000 replicates). ACPRg was log-transformed for modeling. CIR30, corrected insulin response at 30 min.

Corresponding Pearson correlations between observed and predicted log-transformed ACPRg were 0.685, 0.779, and 0.738, respectively (Fig. 2B; Table 2). The individual observed-versus-predicted relationships were maintained across baseline, 12 months, and 15 months, with sample sizes of 257, 220, and 207, respectively (Fig. 2C–E).

Cross-validation of a fixed final formula does not fully account for potential performance overestimation arising from selection of the formula itself. We therefore performed a more stringent nested 5-fold analysis in which the candidate-selection procedure was repeated independently within each outer training set before evaluation in the held-out outer fold. Nested cross-validated R² values were 0.455 at baseline, 0.585 at 12 months, and 0.533 at 15 months. These values were only modestly lower than those obtained with standard 5-fold validation, indicating that formula selection contributed little additional performance inflation.

Robustness to the definition of the fasting value was also examined because the RISE OGTT protocol included samples at −10 and −5 min but no sample at exactly 0 min. The primary analysis defined fasting glucose and C-peptide as the mean of the two pre-OGTT measurements. Recalculation of OEC using either the −10-min or −5-min measurement alone produced essentially unchanged predictive performance across visits, indicating that the results were not materially dependent on averaging the two fasting samples. Together, these analyses showed that OEC performance was maintained under participant-level cross-validation, remained similar under nested validation, and was insensitive to the definition of the fasting value.

### External validation in the BetaFat Study

Internal validation assesses performance in held-out participants from the same study population, whereas external validation provides a more stringent test of whether the relationship generalizes to an independent cohort. We therefore evaluated OEC in the Beta Cell Restoration Through Fat Mitigation (BetaFat) Study, a separate adult RISE study of participants with impaired glucose tolerance or recently diagnosed type 2 diabetes who underwent OGTT and hyperglycemic clamp assessments at baseline, 12 months, and 24 months. The OEC–ACPRg regression model was calibrated using baseline data from the RISE Adult Medication Study, after which the model coefficients were fixed and applied unchanged to BetaFat. No recalibration or refitting was performed using BetaFat data.

The Adult-derived OEC model retained predictive performance at all three BetaFat assessments. Predictive R² was 0.552 at baseline, 0.414 at 12 months, and 0.389 at 24 months, with corresponding 95% confidence intervals of 0.370–0.715, 0.248–0.567, and 0.207–0.590, respectively (Fig. 3A; Table 3). Pearson correlations between predicted and observed log-transformed ACPRg were 0.763, 0.700, and 0.640 at the corresponding visits (Fig. 3B), with 95% confidence intervals of 0.659–0.862, 0.588–0.804, and 0.538–0.784, respectively. At the individual-participant level, observed and predicted ACPRg values remained closely related at baseline, 12 months, and 24 months in BetaFat (Fig. 3C–E). Performance was highest at baseline and attenuated at follow-up, but predictive R² remained positive and the observed–predicted correlations remained substantial at both follow-up assessments despite the absence of model refitting. Together, these results show that OEC retained predictive performance when the model derived in the RISE Adult Medication Study was applied without recalibration to the independent BetaFat cohort.

**Figure 3.**
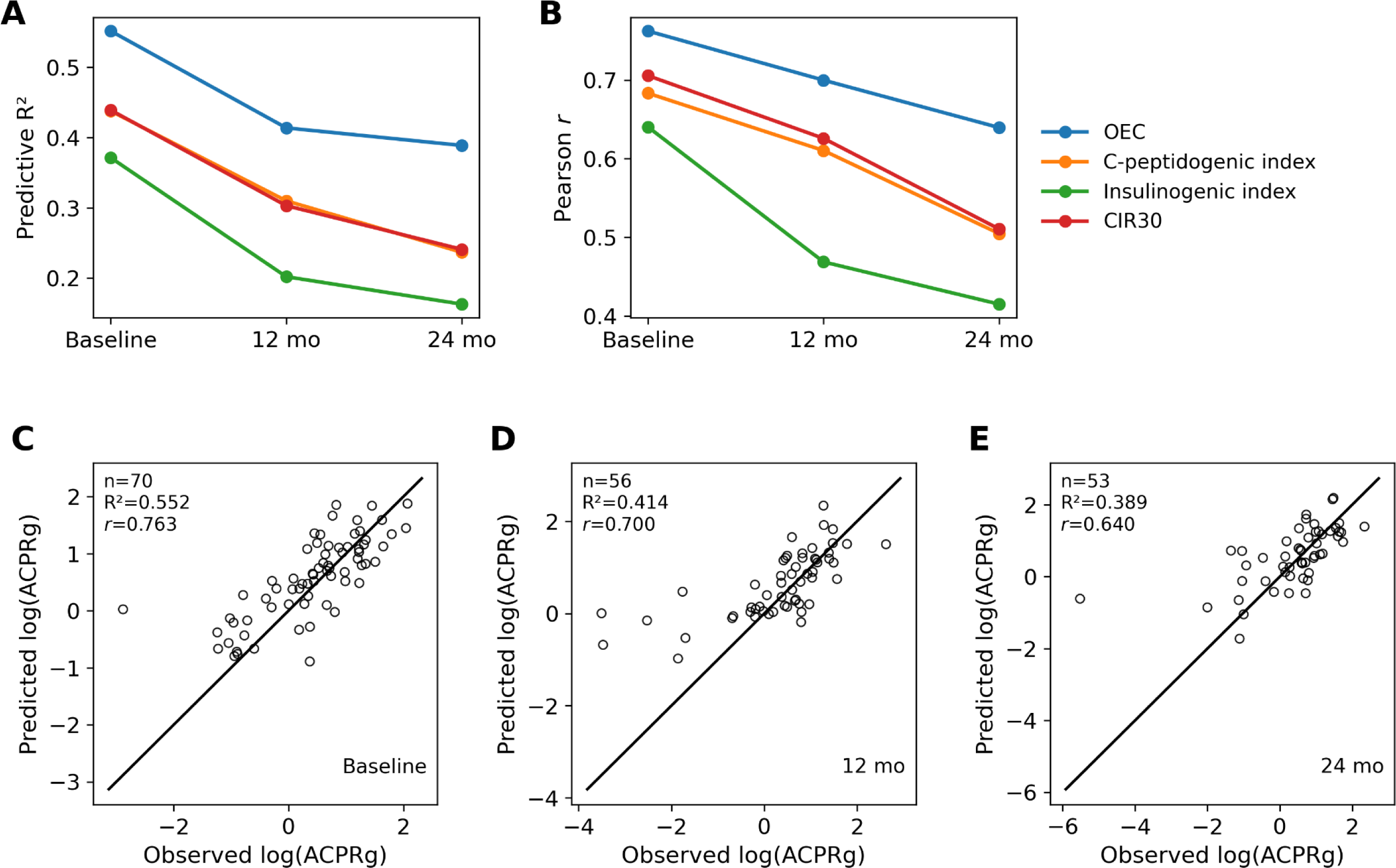
External validation of OEC in the Beta Cell Restoration Through Fat Mitigation (BetaFat) Study. Predictive R² **(A)** and Pearson r **(B)** for OEC, the C-peptidogenic index, insulinogenic index, and CIR30 at baseline, 12 months, and 24 months. Calibration models were derived from baseline data in the RISE Adult Medication Study, fixed, and applied to BetaFat without recalibration or refitting. Observed and Adult-derived predicted log(ACPRg) values for OEC are shown at baseline **(C)**, 12 months **(D)**, and 24 months **(E)**. Individual participants are shown as open circles; the diagonal line represents identity. Sample size, R², and Pearson r are reported in each panel.

**Table 3.** External validation of OEC and conventional OGTT indices in the Beta Cell Restoration Through Fat Mitigation (BetaFat) Study.

| Visit | n | Index | Predictive R<sup>2</sup> (95% CI) | Pearson r (95% CI) |
| --- | --- | --- | --- | --- |
| Baseline | 70 | **OEC** | **0.552 (0.370 - 0.715)** | **0.763 (0.659 - 0.862)** |
|  |  | C-peptidogenic index | 0.438 (0.202 - 0.658) | 0.683 (0.511 - 0.857) |
|  |  | Insulinogenic index | 0.371 (0.168 - 0.562) | 0.640 (0.477 - 0.796) |
|  |  | CIR30 | 0.439 (0.273 - 0.601) | 0.706 (0.591 - 0.822) |
| 12 months | 56 | **OEC** | **0.414 (0.248 - 0.567)** | **0.700 (0.588 - 0.804)** |
|  |  | C-peptidogenic index | 0.310 (0.127 - 0.462) | 0.610 (0.471 - 0.741) |
|  |  | Insulinogenic index | 0.202 (-0.066 - 0.395) | 0.469 (0.261 - 0.682) |
|  |  | CIR30 | 0.303 (0.154 - 0.452) | 0.626 (0.495 - 0.746) |
| 24 months | 53 | **OEC** | **0.389 (0.207 - 0.590)** | **0.640 (0.538 - 0.784)** |
|  |  | C-peptidogenic index | 0.237 (0.062 - 0.462) | 0.505 (0.385 - 0.707) |
|  |  | Insulinogenic index | 0.163 (-0.011 - 0.423) | 0.415 (0.237 - 0.672) |
|  |  | CIR30 | 0.241 (0.069 - 0.471) | 0.511 (0.359 - 0.706) |

Note: Prediction models for each index were derived from baseline data in the RISE Adult Medication Study, fixed, and applied to BetaFat without recalibration or model refitting. Predictive R<sup>2</sup> and Pearson r were calculated from the resulting external predictions. The same participants were used for all indices within each visit. 95% confidence intervals were estimated by participant-level bootstrap resampling (10,000 replicates). ACPRg was log-transformed for modeling. CIR30, corrected insulin response at 30 min.

### Comparison with conventional OGTT indices

We next examined whether the predictive performance of OEC exceeded that of established OGTT-derived measures of early β-cell response. OEC was compared with the C-peptidogenic index, insulinogenic index, and corrected insulin response at 30 min (CIR30), with all indices evaluated in the same complete-case participants within each visit.

In the RISE Adult Medication Study, OEC consistently showed the highest cross-validated predictive performance. Predictive R² values for OEC were 0.469, 0.607, and 0.544 at baseline, 12 months, and 15 months, compared with 0.361, 0.437, and 0.445 for the best-performing conventional index, the C-peptidogenic index (Fig. 2A; Table 2). The same pattern was observed for Pearson correlation: OEC yielded r values of 0.685, 0.779, and 0.738, compared with 0.601, 0.661, and 0.667 for the C-peptidogenic index (Fig. 2B). The insulinogenic index and CIR30 showed lower predictive R² and correlations at each visit.

The advantage of OEC was maintained in the independent BetaFat validation. Using models derived from the RISE Adult Medication Study and applied without refitting, OEC achieved predictive R² values of 0.552, 0.414, and 0.389 at baseline, 12 months, and 24 months, respectively, whereas the highest corresponding values among the conventional indices were 0.439, 0.310, and 0.241 (Fig. 3A; Table 3). Pearson correlations for OEC were 0.763, 0.700, and 0.640, compared with 0.706, 0.626, and 0.511 for the best-performing conventional comparator at the corresponding visits (Fig. 3B). Together, these results suggest that OEC provided consistently greater predictive performance than the conventional OGTT indices across both internal and external validation.

## Discussion

Acute glucose-stimulated β-cell secretion is important in dysglycemia, but the hyperglycemic clamp is impractical for assessing it at scale. Here, we developed OEC to estimate ACPRg from OGTT data and evaluated it in RISE Adult and the independent BetaFat cohort. In RISE Adult, cross-validated R² was 0.47 at baseline, 0.61 at 12 months, and 0.54 at 15 months. In BetaFat, applying the RISE baseline prediction mapping without refitting yielded R² values of 0.55 at baseline, 0.41 at 12 months, and 0.39 at 24 months. Nested cross-validation and sensitivity analyses of the definition of fasting values supported the robustness of these findings.

The OEC Index was designed around three physiological considerations. First, the 100/G₀ term accounts for fasting glycemia, which reflects β-cell sensitivity to glucose in the fasting range and influences the response to a subsequent glucose load. Second, the geometric mean of incremental and total glucose AUCs captures both the glucose rise above fasting and the total glucose load experienced by the β-cell. Third, OEC uses the first 60 min, aligning its window with the acute response represented by ACPRg. The index contains no fitted regression coefficients, retaining a simple and physiologically interpretable structure.

The present study has several strengths. Both cohorts included paired OGTT and clamp measurements at multiple visits, allowing OEC to be evaluated directly against ACPRg. Internal validation used participant-level fivefold cross-validation, with all observations from each participant retained in the same fold to prevent information leakage. Nested cross-validation, in which index selection was repeated within each outer training fold, showed only modest attenuation relative to standard cross-validation, indicating that the performance of OEC was not materially inflated by formula selection. The RISE baseline model was then applied to the independent BetaFat cohort without refitting or recalibration, providing a stringent assessment of transportability. Results were consistent across sensitivity analyses examining the definition of fasting values and the length of the response window. Finally, the OEC formula contains no empirically fitted coefficients and requires only standard OGTT glucose and C-peptide measurements, making it transparent, reproducible, and straightforward to calculate.

Several limitations should be acknowledged. First, the BetaFat cohort was relatively small, with 70 participants at baseline and 53 at 24 months, resulting in wide confidence intervals around estimates of predictive performance. Performance was also strongest at baseline and attenuated at follow-up; the reasons for this attenuation are uncertain and may reflect differences in cohort characteristics, intervention exposure, or other longitudinal changes. Second, both cohorts consisted of adults with impaired glucose tolerance or recently diagnosed type 2 diabetes, and whether OEC performs similarly in normoglycemic individuals, youth, people with longer-duration diabetes, or more diverse racial and ethnic populations remains unknown. Third, OEC was developed using the RISE OGTT protocol, which included fasting samples at −10 and −5 min and measurements at 30 and 60 min, and its performance with alternative sampling schedules has not been established. Finally, although clamp-derived ACPRg provides a rigorous physiological reference measure of acute β-cell secretory capacity, it represents one construct of β-cell function, and validation against complementary measures such as IVGTT-derived responses was not performed.

These findings have several implications. By providing an OGTT-based approximation of acute β-cell secretory capacity from glucose and C-peptide measurements, OEC may extend physiological assessment of early β-cell function to settings in which hyperglycemic clamps are impractical. This could be particularly useful in large observational studies, clinical trials, and existing cohorts with archived OGTT data, enabling broader investigation of the determinants, progression, and treatment responsiveness of β-cell dysfunction. Before wider application, however, the frozen OEC formula should be evaluated in larger and more diverse populations, across different OGTT sampling schedules and C-peptide assays, and for reproducibility and longitudinal responsiveness. Future studies should also determine whether OEC provides prognostic or treatment-response information beyond conventional glycemic measures and established OGTT indices. Where an estimate expressed in clamp ACPRg units is desired, separately calibrated mappings from OEC to ACPRg could also be developed and validated.

## Conclusion

In conclusion, the OEC Index provides a simple OGTT-based measure that captures acute β-cell secretory capacity as assessed by clamp-derived ACPRg. Its performance across internal and external validation supports the feasibility of estimating this physiological construct from routinely obtainable glucose and C-peptide measurements. With further validation in broader populations, OEC may therefore provide a practical tool for extending assessment of early β-cell function to larger clinical and research settings in which hyperglycemic clamps are not feasible.

## Statements and Declarations

### Ethical approval

The present study was approved by the Institutional Review Board of Wayne State University.

### Consent to Participate

Not applicable.

### Author Contributions

R.Z. conceptualized, designed the study, analyzed the data, and wrote the paper.

### Ethical Responsibilities of Authors

The author has read, understood, and complied, as applicable, with the statement on Ethical Responsibilities of Authors in the Instructions for Authors.

## Data Availability

The participant-level data from the RISE Adult Medication Study and the Beta Cell Restoration Through Fat Mitigation (BetaFat) Study analyzed in this study were obtained from the NIDDK Central Repository and are available through the repository subject to its data-access procedures and requirements.

https://repository.niddk.nih.gov/home

